# Lessons in Implementing Complex Interventions in a Public Health Emergency: A Process Evaluation of the California Contact Tracing Support Initiative

**DOI:** 10.64898/2026.02.07.26345668

**Authors:** Erica N. Rosser, Melissa A. Marx, Soim Park, Lana Feras Aldos, Riaa Dutta, Kyra H. Grantz, Kyu Han Lee, Laura-Marie Peeples, Emily S. Gurley, Elizabeth C. Lee

## Abstract

**Background:** Emerging in January 2020, the SARS-CoV-2 pandemic quickly exposed the limitations of traditional contact tracing and overwhelmed the contact tracing efforts of US health departments. In response, Kaiser Permanente partnered with the Public Health Institute to launch the California Contact Tracing Support Initiative. This innovative, clinically integrated program aimed to link Kaiser Permanente members diagnosed at their facilities directly with contact tracing and supportive clinical care via their network. This approach promised to address key logistical and behavioral challenges hampering traditional public health agencies. This paper evaluates the program’s implementation in two California counties.

**Methods:** We conducted a retrospective, mixed-methods process evaluation of program activities from August 2020 to June 2021, including contact tracing implementation in Fresno and San Bernardino Counties. Our methods included scoping discussions with program stakeholders, development of an epidemiological timeline and program impact model, and document review. We also conducted semi-structured interviews with program stakeholders and staff. Interviews were conducted and audio-recorded via Zoom, transcribed, and analyzed in NVivo using inductive and deductive coding with a Framework Approach.

**Results:** We reviewed 474 program documents and interviewed 47 participants. Study findings highlighted difficulties in adapting program scope due to competing partner visions of program mission and collaboration. Unforeseen data demands and complex external data sharing with public health systems further complicated and delayed program implementation.

**Conclusion:** Evaluation of this contact tracing program offers key insights into public health interventions during emergencies. While the California Contact Tracing Support Initiative’s integrated design showed promise, challenges arose from data systems, inter-organizational dynamics, and planning. Findings emphasize the need for clear operational steps, real-time data monitoring, defined roles, and formalized public-private partnerships in preparedness planning. These are key lessons for future complex public health interventions, especially regarding adapting programs versus maintaining fidelity amidst evolving contexts.

## INTRODUCTION

In January 2020, the SARS-CoV2 pandemic was documented for the first time in the United States. The pandemic spread rapidly and has, as of May 2025, caused over 750 million infections and 7 million deaths globally, including 1.2 million deaths in the US^1^. When the pandemic began, only non-pharmaceutical interventions, including case investigation and contact tracing (“contact tracing”), were available to prevent transmission^2^. While tracing contacts, investigating and isolating cases, and identifying and quarantining case’s close contacts, has been used to prevent transmission of infectious diseases for centuries, effective contact tracing is time- and personnel-intensive, and requires specially trained interviewers^3–5^.

For acute infections like COVID-19, which have a relatively short incubation period, the effectiveness of contact tracing quickly diminishes unless there are minimal delays in notifying and isolating both index cases and their contacts. Delays from symptom onset to seeking care and testing, laboratory test turnaround, and the process of contacting cases and contacts affect the timeliness of contact tracing^6–8^. Contact tracing effectiveness during the COVID-19 pandemic was also limited by considerable behavioral challenges, including low response rates to public health calls, reluctance to disclose information due to mistrust in government, public health and digital technology, and fear of stigma or imposing quarantine restrictions on others^8–11^. A survey of over 10,000 US adults conducted in July 2020 found that 41% were unlikely to speak with a public health official about COVID-19 by telephone or text message, and 27% were uncomfortable disclosing contacts^11^. These challenges were further corroborated by a 2020 multi-state cross-sectional study where contact tracing efforts reportedly yielded no contacts for two-thirds of COVID-19-positive individuals either because they could not be reached for an interview or, if interviewed, did not disclose contacts^8^.

Dramatic surges in new COVID-19 cases overwhelmed contact tracing capabilities at health departments^3–5^, including in California, where county health departments struggled to keep up with contact tracing demands during a surge in cases that occurred after stay-at-home orders were lifted in May 2020^12^. In August 2020, Kaiser Permanente (KP), an integrated health maintenance organization (HMO), launched the California Contact Tracing Support Initiative (CCTSI). This innovative collaboration, led by the Public Health Institute (PHI), a public health implementation organization, aimed to support pandemic response with contact tracing for KP members who tested positive for COVID-19 (i.e., cases) and their contacts across seven California counties. The central set of activities under CCTSI was designed to connect KP members diagnosed within KP facilities with integrated contact tracing, supportive services, and clinical care provided through KP’s own network operating in different counties.

This novel, clinically integrated approach was designed to address the pervasive delays and challenges plaguing traditional contact tracing efforts during the pandemic. By leveraging their existing infrastructure, KP wanted to link their laboratory test results with their own contact tracing teams. This integration aimed to significantly reduce crucial time lags in the contact tracing process. Establishing seamless information flow from diagnosis within the HMO to immediate contact tracing could enable the program to drastically reduce isolation and quarantine times — a critical advantage given COVID-19’s short incubation period. Moreover, this model aimed to capitalize on the pre-existing relationship between KP members and their clinicians, which was perceived to have fostered a trust often absent in interactions with traditional public health agencies.

As part of the program activities, CCTSI staffed and trained “micro-teams” comprised of investigators responsible for contact tracing calls, resource coordinators who connected beneficiaries to support services for isolation and quarantine, and supervisors. These micro-teams were structured for nimble and rapid deployment across jurisdictions ^13^. PHI also built a data system to capture contact tracing data for KP members who tested positive for COVID-19 at KP facilities and to monitor program progress (See full description of program and personnel roles in Annex 1).

KP commissioned an evaluation of this contact tracing initiative, which was noteworthy because programs of this type had not been evaluated previously. We conducted a process evaluation of CCTSI’s clinically integrated contact tracing approach in two California counties, including the initial design and its subsequent evolution in the context of early pandemic emergency response.

## METHODS

We conducted a retrospective, mixed-methods evaluation of CCTSI hiring, training, and contact tracing activities from August 2020 to June 2021, including program implementation in Fresno and San Bernardino Counties. We employed a suite of qualitative methods, and synthesized findings to document the program design and theory of change in an impact model. We also developed timelines to describe CCTSI activities in the broader epidemiological context.

### QUALITATIVE METHODS

To scope the program, we compiled key documents provided by PHI and KP, including meeting minutes, training materials, contracts, and design plans. Document review helped us to pinpoint internal and external factors impacting the program. After summarizing documents, we addressed clarifying questions to PHI and KP leadership and requested supplementary documents when necessary. To develop a full understanding of the program’s intended design, implementation, and the experiences of staff and partners, we conducted semi-structured key informant interviews with stakeholders and in-depth interviews with staff. Interviews were guided by questions aimed at eliciting insights into the program’s design and execution, and the perspectives of those involved.

We carefully selected interview participants by reviewing roles and responsibilities identified in program documents and through initial discussions with PHI and KP. This process helped us identify individuals who were highly knowledgeable about different aspects of the program. We aimed for an equal number of stakeholders from both KP and PHI. We also interviewed program staff from each of the micro-team roles. PHI and KP leadership supported recruitment by notifying potential participants via staff meetings and email that our evaluation team might contact them for interviews, and by inviting them to express interest in participating in the study by contacting the evaluation team. Snowball sampling was also deployed during interviews with participants suggesting additional interview candidates. Sample size was guided by the principle of data saturation, whereby interviews continued until no new significant themes or perspectives emerged.

All interviews were conducted via password-protected Zoom calls and audio-recorded except two interviews in which participants declined consent to recording, and we used expanded field notes to document the conversations. Audio recordings were transcribed verbatim, and transcripts were analyzed using NVivo data analysis software (QSR International, Melbourne, Australia). We employed inductive and deductive approaches to develop the codebook used for all interview transcripts. We used a Framework Approach to analyze the interview data, enabling systematic coding and thematic analysis of the qualitative insights gathered. ^9,10^

Participants provided oral informed consent and were offered $25 for their participation. Ethical approval for this study was granted by the Johns Hopkins Bloomberg School of Public Health Institutional Review Board (IRB No. 17623).

### IMPACT MODEL

Informed by the qualitative methods described above, we developed a theory of change for the CCTSI program. We then constructed an impact model to depict the causal chain that links program inputs, activities, and outputs with broader public health outcomes and program impact.

### PROJECT TIMELINE IN CONTEXT

COVID-19 case data were extracted from the Johns Hopkins University COVID-19 database^14^ to construct a timeline of weekly COVID-19 cases in California from January 2020 to September 2021. We juxtaposed this epidemiologic timeline with project milestones.

## RESULTS

### SUMMARY OF DATA COLLECTED

#### Document review

We reviewed a total of 474 inventoried files on topics that included staff training and development (N=136), KP-PHI meeting agendas and notes (N=130), program implementation (N=45), weekly updates sent to KP (N=44), staff recruitment (N=40), workforce development (N=32), communications (N=23), and data dictionaries, grant and contract documents, data procedures, progress reports, and personnel directories and organizational charts (N<7 each) (Supplemental **Error! Reference source not found**.).

#### Qualitative interviews

Of 100 people contacted for interviews, 47 individuals were interviewed (Supplemental **Error! Reference source not found**.). PHI staff represented about half of key informant interviews (N=13); other informants included KP staff (N=9) and representatives from the California Health Medical Reserve Corps (N=1) and the San Bernardino County Health department (N=1). In-depth interviews were conducted with CCTSI contact tracers (N=12), resource coordinators (N=4), supervisors (N=5) and team managers (N=2).

### IMPACT MODEL

### PROGRAM IMPLEMENTATION

#### Epidemiologic Timeline

The CCTSI program was announced during the first spike in cases in California. The San Bernardino site was launched just before the period subsequently known as the Delta variant wave in the winter of 2020, and the Fresno site was launched in the middle of the Delta variant peak (Figure 2).

**Figure 1.**
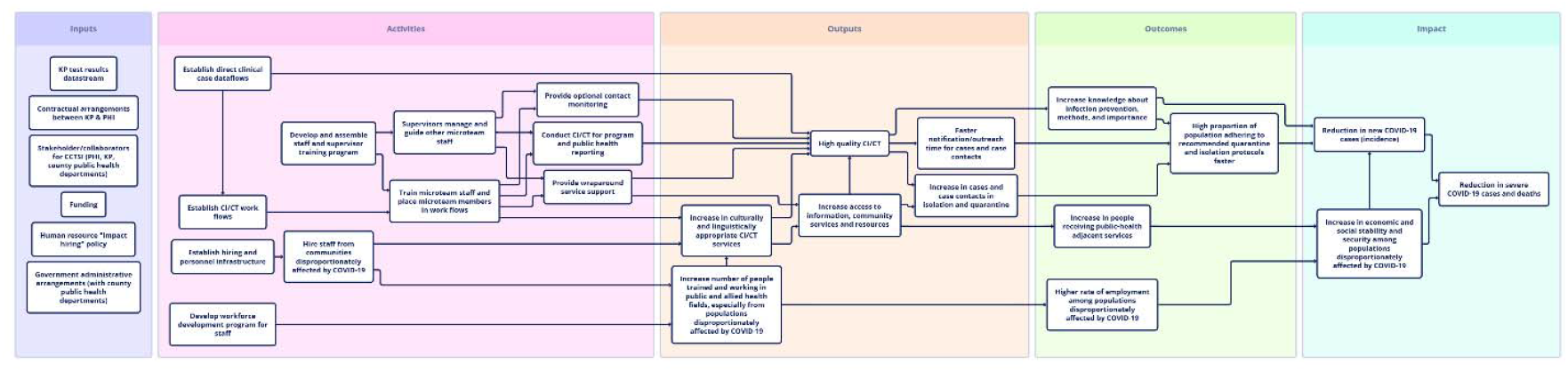
Impact model for the CCTSI program. This diagram shows how program inputs (lavender) and case investigation and contact tracing (CI/CT) activities (pink) link to measurable program outputs (orange), which drive program health and employment outcomes (green) and impact (blue).

**Figure 2.**
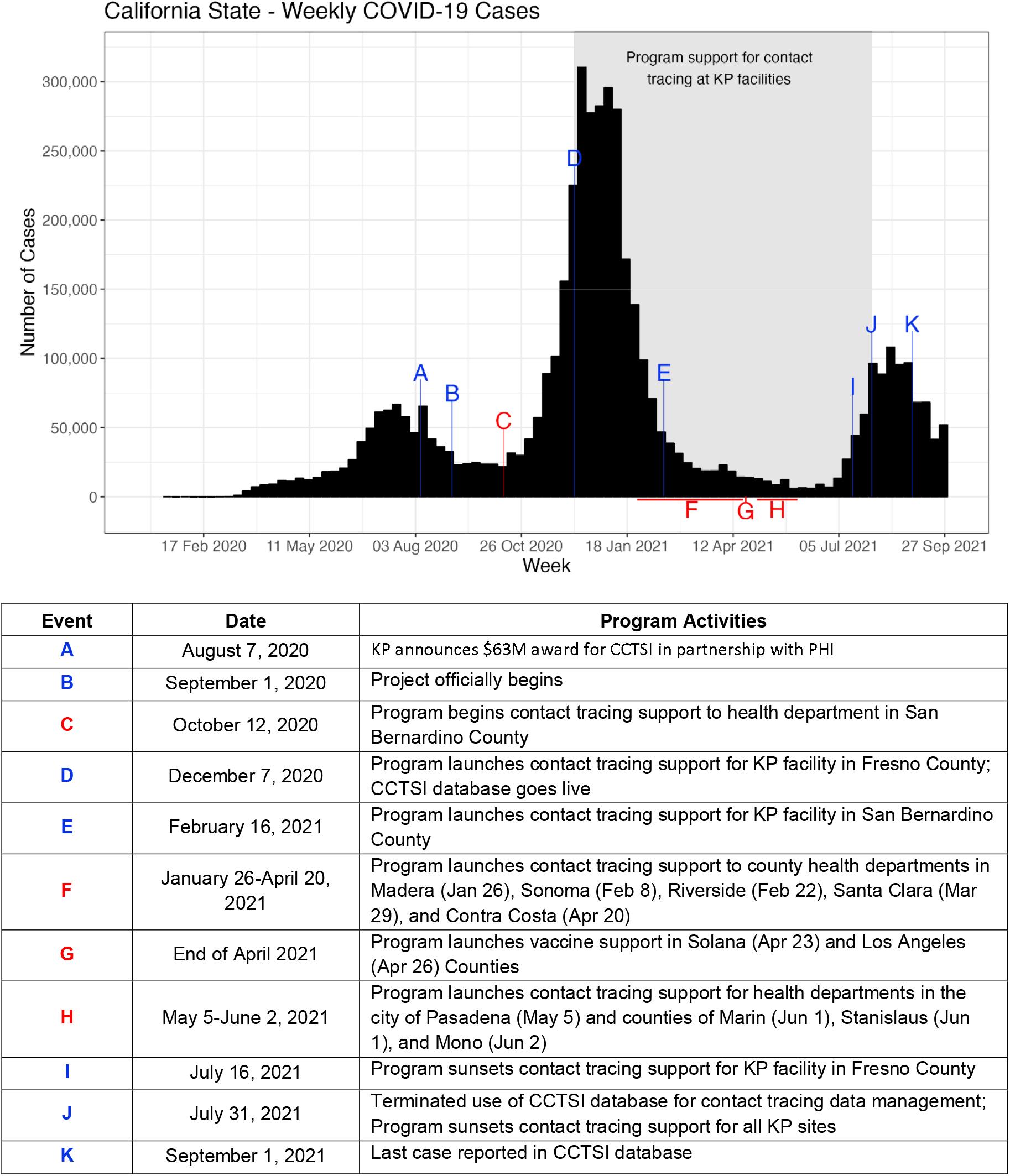
Timeline of weekly COVID-19 cases in California (January 2020 to September 2021) annotated with select project milestones. Blue annotations highlight events evaluated in this manuscript, while red annotations label other select CCTSI program activities. The period highlighted in grey indicates the project period for CCTSI contact tracing activities. COVID-19 case counts were obtained from the public Johns Hopkins University Center for Systems Science and Engineering database.

#### Building the KP-PHI Partnership

With a backdrop of a challenging pandemic context and novel partnership, the process evaluation found that although many CCTSI activities were implemented with high fidelity to the initial program vision, there were implementation challenges related to data and stakeholder alignment.

### Inter-Organizational Work Dynamics

Despite shared goals and good intentions, KP’s and PHI’s perspectives on collaboration faced significant challenges due to differences in program vision and a “*clash of cultures*”. The immense complexity of the endeavor, coupled with the urgent nature of the public health emergency created an environment where expectations on roles and engagement often conflicted. While KP expected PHI to lead implementation, they also wanted to take an active role in day-to-day operations and decision-making. Accustomed to the rapid feedback of a clinical setting, KP prioritized patient experience within CCTSI and aimed for an integrated contact tracing model tightly aligned with their clinical practices. Conversely, PHI stakeholders were accustomed to relative autonomy in their grant-funded research and some perceived KP’s “hands-on” approach as disruptive to program execution. One PHI stakeholder remarked, “Kaiser did not treat this as a grant, they treated it as a contract. And they were involved in every minor detail…It was, ‘The money is here. We wanna tell you how to do it, when to do it, who to engage with, how to engage and be part of the entire process.’”

KP stakeholders explained that their executive sponsorship felt a responsibility to ensure the CCTSI was successful due to the perceived “*high stakes*” of the initiative. As a closed health system, granting an external organization access to patient data was a significant move, and the grant itself was one of the largest KP had ever provided. One KP stakeholder reflected that internal stakeholder requests for “*real time data about how the program was performing*” created a “*fishbowl*” environment, and that the partnership with PHI would have been more effective had they been able to troubleshoot issues in smaller working groups, rather than hearing about new developments for the first time in large meetings with 20 people. This same stakeholder, who described an early “*hand in glove*” working relationship with PHI, went on to explain that collaboration also became more complicated as program workstreams developed. A PHI stakeholder echoed this sentiment saying, *“I think this was a complex endeavor, and one where there were lots of forces…And it was a little bit like the blind men and the elephant that from one perspective. You thought you understood what it was, but somebody was at the other end of the elephant, and they had a different perspective. Everybody wanted to take good care of the elephant*.*”*

Across organizations, stakeholders acknowledged the immense effort invested in aligning themselves but also agreed that, navigating the urgency of a public health emergency amid remote work left little room for addressing underlying differences in expectations and collaboration styles. A PHI stakeholder reasoned, “*Some of the time tested and proven approaches to project management, such as pausing for a moment to make sure everybody understands the scope, making sure we’re all agreeing about who’s doing what, [what] our objective is, who has what roles, what lanes, what decision-making authorities, outlining a work plan, being clear about what decisions need to be taken*… *I think those were shelved in the name of the crisis*.”

#### Unforeseen Data Demands Hampered Implementation and Site Expansion

Establishing contact tracing monitoring and performance procedures emerged as a crucial, yet insufficiently anticipated CCTSI activity. Although KP-PHI contracts stipulated reporting on contact tracing performance indicators, they lacked specifics on metrics, quantitative benchmarks, and reporting frequency. This allowed for differing interpretations, ultimately revealing a fundamental misalignment between PHI’s assumption of the periodic reporting typical in grantor-grantee relationships, and KP’s expectation for on-demand performance data. This discrepancy in expectations resulted in unanticipated work for PHI, with one PHI stakeholder recounting, “*While [PHI] were busy trying to staff up, get everything running, we had people on the KP side who wanted constant updates on how we were doing, how it was working, what we needed to improve*.*”* The unanticipated volume and urgency of these requests did not allow time for sufficient data cleaning and management. As described by another PHI stakeholder, *“The first four to six weeks of data… were terrible. Some of that is because we [PHI] never planned to create these reports, and we hadn’t done any quality control checks on our data collecting. So, numbers didn’t match, totals didn’t match*.*”* Discrepancies in reported data eroded KP stakeholder confidence in program data quality and scalability. A KP stakeholder explained:

> *“I think from the PHI side, they felt like we were holding them back from scaling it up fast enough. And from the KP side, we felt like they weren’t giving us the data that we needed to confidently scale it up fast enough. So it was like this feeling that we just weren’t moving quickly enough on both of our sides, but we both intuited different reasons for that*.*”*

PHI rapidly hired staff to meet grant hiring and training deadlines, but KP was hesitant to expand contact tracing support to new sites while data quality remained poor. By mid-January 2021, COVID-19 case counts were climbing in California, and although trained CCTSI staff were available to meet the need, teams were not deployed to new sites, leading to a drop in staff morale. One PHI stakeholder noted, *“[CCTSI] Staff saw those [high case] numbers and that was another morale issue – ‘Oh, I know there’s thousands of cases there today, but we’re only going to do these 30 and [since] we have so many people, everybody gets one’*.*”* Matching staff levels with fluctuating caseloads to optimize staff placement across counties remained a constant challenge, with a PHI stakeholder likening staffing deployment to “*turning the Titanic*.” Difficulty in understanding reported contact tracing metrics, coupled with PHI’s emphasis on meeting staffing objectives caused frustration among some KP stakeholders with one stating, *“PHI kept saying, “We’ve hired this many contact tracers” … So, it kind of seemed to us like hiring was their big feat*.*”*

#### Competing Program Visions Led to Divergent Perceptions of Program Adaptations

The grant agreement establishing CCTSI indicated that the program would provide “contact tracing support services and other science and evidence based public health prevention activities and practices,” suggesting that adaptations might be expected as the pandemic evolved. In practice, KP and PHI interpreted this principle differently, disagreeing on when and how to adapt the program. PHI viewed the program’s scope as dynamic and adaptable as public health needs evolved, *“Because it [CCTSI program] wasn’t just contact tracing. My understanding is that it was supposed to be a response to COVID-19. And pandemics don’t stay in their box. So, in order to fight it, I don’t think we can fight it in our box*.*”* PHI viewed deviations from the envisioned CCTSI model — such as shifting program staff to provide direct support for county contact tracing and vaccine outreach —as chances to address critical needs. PHI stakeholders expressed disappointment, even “*heartbreak*,” at missed opportunities to expand activities in response to waves of cases and county requests:

> *“The way the grant was constructed is a brilliant way to implement in a pandemic because it has maximum flexibility which you would need in order to go with the flow and meet the moment. …I can’t speak for [KP], but in my opinion, that is not what [KP] was looking for. But, that rigid and strict control framework … It doesn’t work well in a pandemic scenario because you will, as we did, always miss the moment*.*”*

KP stakeholders were aware that as the pandemic evolved there would be pressure to respond to emerging needs and sympathized with PHI’s perspective with one saying, *“I can understand why PHI out in the community, hearing all these demands and all these needs that are bubbling up from local health departments, they’d want to adapt, and you’d want to modify, and you’d want to support*.*”* However, KP program architects wanted to maintain focus on the innovation of integrating contact tracing into clinical care, believing it could address the limitations of traditional contact tracing and improve upon the US COVID-19 response. KP therefore attempted to reduce scope creep and continue implementing the original vision approved by their board:

> *“Let’s test [this new contact tracing model], let’s make sure it works. And not be caught up in a lot of the-‘Well, this health department wants to do this, and this other local health department who wants to do this. And this health department wants to do this other model’. And we end up diluting the focus of what those millions of dollars were supposed to be doing*.*”*

### Shifting Priorities: Additional Changes to Program Design

As the program became more established, CCTSI activities did expand beyond the initial scope of contact tracing support to KP healthcare beneficiaries. In the end, a substantial effort went to direct deployment of staff to support contact tracing investigations at county health departments (See events C, F, and H in Figure 2). After a COVID-19 vaccine emerged in December 2020, under-resourced counties began seeking CCTSI support for vaccine outreach efforts. While there was initial resistance from KP leadership to expand the program scope and “*morph some of the grant dollars to focus on* …*outreach for vaccination*,” ultimately CCTSI secured approval in April 2021 to provide vaccine support with up to 5% of the grant budget (See event G in Figure 2).

#### External Partnerships: Political Context and Access to Data Systems

The CCTSI program was designed to provide support to the public health system during a pandemic with one external stakeholder reflecting that the program “*took a lot of the burden off of overtaxed health departments*.” However, the involvement of nongovernmental organizations in delivering a public health service necessitated buy-in and complex coordination across diverse stakeholders, including county and state health departments, the California governor’s office, and KP medical facilities (Figure 3), for contact tracing site selection and to navigate complex public health data and reporting systems.

**Figure 3.**
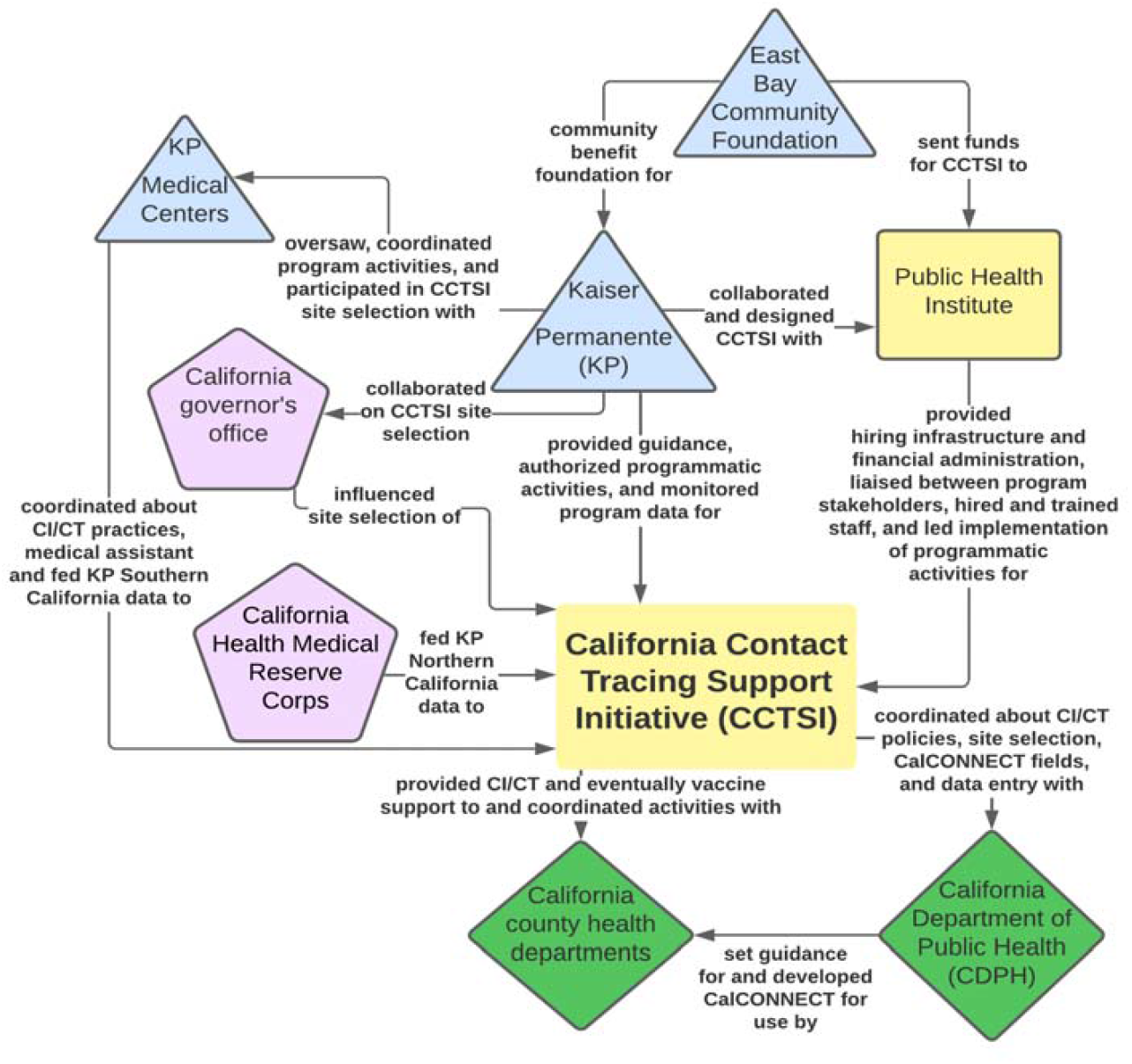
Stakeholder landscape in CCTSI implementation. Blue triangles indicate KP-affiliated organizations, yellow rectangles PHI-affiliated. Green diamonds show broader public health stakeholders. Pink pentagons show stakeholders outside the program and California public health actors.

Stakeholders remarked that beyond its innovative contact tracing approach, CCTSI uniquely addressed the challenge of providing private support to a public health system in crisis, with one PHI stakeholder saying,

> *“It kind of begs the question, what role do all of these institutions play in this pandemic long run? What are the right lanes for people? What is Kaiser’s lane? What’s PHI’s lane? What’s the lane of the non-profit? What’s the lane of the medical institution? What’s the lane of the state or the county? We’re a year into this, but I think those are the questions that we didn’t necessarily ask a year ago because we were just trying to help. But I think that is something that I think about with the clinically integrated model*.*”*

Initial program discussions prioritized county health department and stakeholder engagement and script development, yet stakeholders later reflected that understanding the full scope of data exchange challenges (described below) ultimately required several months of unrecognized, additional dedicated effort. Navigating the political context was also critical to the data exchange issue.

In brief, CCTSI staff needed access to COVID-19 data feeds from testing facilities to perform contact tracing in a timely manner. However, they were unable to directly access electronic health records (EHRs) in KP systems, so they sought alternative options for accessing laboratory confirmation data. At the same time, data exchange between CCTSI and county public health systems was crucial for avoiding duplicate case investigations and ensuring proper reporting back to county health departments for public health surveillance. However, stakeholders reported that negotiating access to CalCONNECT, California state’s contact tracing data system, proved unexpectedly challenging due to various political considerations. Without state-level agreement to share data, PHI had to negotiate access to CalCONNECT on a county-by-county basis. To avoid delays, in September 2020, PHI developed a custom CCTSI database to accept case data feeds. A micro-team member reflected:

> *“I had the experience in the county where labs [COVID-19 confirmatory tests] were coming in five, seven days after the person was already diagnosed. To have a system like [the CCTSI database], where you got them in 24, 48 hours, for us – we were making an impact in real-time because in the county it was so backed up that it by the time we called these people, either they were outside of their isolation [period], or we were getting them towards the end of their isolation*.*”*

This database also paved the way for more efficient program implementation, because it supported CCTSI-specific workflows and facilitated collecting additional data for program evaluation.

Further complicating matters, federal guidance on contact tracing data standards and software were lacking, and CalCONNECT was itself under development and not universally adopted by counties. CCTSI database developers faced constant challenges with maintaining data system interoperability, and CCTSI staff were required to perform manual double data entry into the CCTSI database and CalCONNECT.

To implement the program quickly in response to the winter 2020 surge, PHI partnered with the California Health Medical Reserve Corps to establish standardized data feeds for Northern California KP facilities. Operational differences between KP’s regional systems prevented a similar arrangement for Southern California KP facilities so CCTSI developed contracts and data acquisition protocols with individual counties and facilities, which proved cost- and time-intensive. Initially, stakeholders reported faster case data import into the CCTSI database compared to county health department systems, but by summer 2021 CalCONNECT import speeds improved, and it was used in place of the CCTSI database.

## DISCUSSION

While the CCTSI succeeded in testing a clinically integrated contact tracing approach and scaling support to multiple California counties, it also encountered setbacks related to data systems, inter-organizational dynamics, and program planning. Despite the shared goal to support contact tracing in California, the KP-PHI partnership faced challenges due to divergent expectations of leadership, engagement levels, and operational autonomy. The program also faced delays in both implementation and expansion, due to factors such as lack of clear reporting expectations, a complicated political context, and challenges in establishing data exchange.

The updated Medical Research Council (MRC) guidance on evaluating complex interventions emphasizes that complex interventions are dynamic and interact necessarily with existing structures and processes, which means that ‘context’ is a key determinant of intervention success^15^. We employed the MRC framework when interpreting our evaluation findings, as acknowledging the urgency of COVID-19 pandemic response was critical for understanding program implementation decisions. The pandemic context of the CCTSI program led to the establishment of a new partnership between PHI and KP and innovative program design that sought to resolve observed contact tracing challenges. It also strongly contributed to diverging stakeholder perceptions on whether proposed adaptations away from clinically integrated contact tracing activities represented implementation improvements under the program mission or deviations from the original program design that compromised fidelity. This clash of perspectives among stakeholders reappeared at several decision points in program implementation, from scaling up contact tracing to new KP facilities to expanding activities beyond contact tracing.

The MRC guidance on evaluating complex interventions urges evaluators to develop a program theory of change to document the causal links between program activities and impact at the outset of an evaluation^15^. Solidifying a joint understanding of the program mission and theory of change may have resolved some of these inter-organizational challenges, but it was not a high priority for stakeholders due to the overwhelming urgency to make progress on implementation. We represented the CCTSI theory of change with an impact model and found that CCTSI sought to improve the quality of contract tracing through integration with KP clinical systems and the timeliness of contact tracing by establishing direct data exchange links on COVID-19 confirmatory tests. The impact model also illuminated that the unforeseen inability of KP to share EHRs with CCTSI precipitated a sequence of challenges that delayed implementation, including building and maintaining a custom CCTSI database and identifying other laboratory data streams.

In placing CCTSI in the broader pandemic emergency response context, this evaluation crystallizes key recommendations for program implementers, decision makers, and researchers seeking to design and implement successful programs during public health emergencies. Below, we present these recommendations, which pertain particularly to building new partnerships and navigating complex stakeholder landscapes.

### Recommendation 1: Even during emergencies, implementers of complex programs should invest time to articulate a clear program vision and implementation plan

While program implementation often takes precedence in emergency contexts, it is critical for stakeholders to take time at the program outset to jointly agree on the program mission and theory of change, which includes explicit operational program activities, and roles and responsibilities of involved actors. The CCTSI program’s flexibility could be considered a strength, as it enabled large scale adaptations as the pandemic evolved. However, it also contributed to different definitions of program success and conflicting perspectives on whether adaptations represented program improvements or deviations from the original mission. Had there been a theory of change documented at the outset, establishing data exchange between KP and PHI and between CCTSI and local health departments may have been identified as a core program activity and tackled more proactively.

Even if program scope is expected to change over time, it is helpful to set early expectations with written documentation, which then opens the possibility for re-evaluation as the situation evolves. It may also be fruitful to designate milestones or time points during the program period for re-evaluation or identify a neutral party that can moderate complex relationships.

### Recommendation 2: Monitoring and evaluation of program data in real-time should be established as a core activity when planning complex programs during emergencies

Emergency contexts are inherently dynamic, and it is foreseeable that most emergency response programs would evolve as the situation changes. As such, real-time monitoring and evaluation of program effectiveness should be a core tenet of any such program with dedicated resources and early attention to minimize implementation disruptions. Key monitoring and evaluation activities include the need to proactively identify measurable program metrics, establish a regular reporting schedule, and document how metrics might be linked to program adaptations. Just as rigorous training is essential for quality assurance, continuous data monitoring is essential to the implementation of high-quality public health interventions^16^.

CCTSI stakeholders acknowledged the poor quality of early contact tracing monitoring data and that decisions to expand the program were delayed due to an inability to clearly describe outputs of the contact tracing intervention. Targets for program metrics (e.g., interview X cases per day, reach X% of contacts within 24 hours of elicitation) should be established collaboratively during the design phase to align expectations, even if these targets are revised at a later stage. Tying program metrics to specific decisions (e.g., when X target is met, expand activities to two more sites) enhances decision making transparency, stakeholder alignment towards common goals, resource allocation, and program effectiveness. In the case of COVID-19 contract tracing, program metrics could have also been used to evaluate whether isolation and quarantine were indeed benefiting public health, which has broader policy and societal implications.

### Recommendation 3: Implementers of complex programs during emergencies should consider how to influence the implementation context to improve chances of program success

Implementers may tend to focus on program-specific levers (e.g., modifying a program activity) to achieve program goals, but given that context is a key determinant to program success, where possible, they should also seek to leverage contextual factors such as political considerations, stakeholder connections, or public opinion in ways that promote program activities.

CCTSI required coordination and buy-in from local and state-level governmental actors, particularly related to aligning contact tracing workflows between CCTSI staff and county contact tracers and exchanging laboratory testing and contact tracing data. Some implementation delays were driven by CCTSI’s need to establish data sharing agreements and procedures on a county-specific basis. Had there been more state-level willingness to intervene in establishing data exchange with non-governmental organizations, or stronger state or federal standards and support for contact tracing practice and evaluation^4,17^, some implementation challenges may have been alleviated. While we cannot know the extent to which implementers could have changed these contextual factors during the COVID-19 emergency, we can anticipate that laying the groundwork for external coordination and data sharing during non-emergency periods would be highly advantageous for future emergency responsee^18^.

Public-private partnerships could also be considered as one mechanism for the rapid and flexible scale-up of complex interventions in an emergency. Private organizations may leverage their relatively rapid and flexible administration, while governmental organizations may contribute access to their far-reaching populations and integrated systems.

## LIMITATIONS

Our retrospective, mixed-methods design carries the inherent risk of recall and social desirability bias. To mitigate this, we systematically employed data triangulation, verifying all qualitative interview data against contemporaneous program documents (e.g., contracts and meeting minutes) and quantitative program performance metrics to ground subjective narratives in objective, verifiable evidence. Furthermore, our interview recruitment strategy deliberately ensured a diverse range of perspectives by including interviewees from high-level partners to front-line staff, allowing us to validate key findings through the analysis of often-differing accounts of the same program activities and decisions. Despite these efforts, our findings remain subject to certain limitations. Due to the reliance on voluntary participation, there may be inherent differences between subjects who opted to participate in interviews and those who were unavailable, unresponsive, or declined. Also, despite assurances of confidentiality, participants may have censored their responses when discussing organizations with whom they remained professionally affiliated.

## CONCLUSION

The successful replication and improvement of future large-scale public health interventions depend on rigorous program evaluations. The evaluation of the CCTSI program offers valuable lessons that extend beyond its specific mandate, illuminating key considerations for future large-scale public health interventions and emergency responses, including pandemics. In the context of an emergency, where stakes are high, public visibility is intense, and significant funding is involved, each partner enters the collaboration under high internal pressure to successfully meet their specific organizational objectives. It is within this complex environment that our evaluation highlights a critical finding: while the urgency of a pandemic demands rapid action, persistent operational challenges often stem from inadequate initial alignment of the shared program vision, fundamental program activities, and expectations among partners. Despite the implementation hurdles faced by the CCTSI, the program’s experience also underscores the potential of public-private partnerships to rapidly mobilize essential surge capacity. Finally, while program flexibility is essential in a dynamic pandemic environment, CCTSI’s experience reveals the importance of establishing clear parameters for adaptability versus program fidelity during initial program design and planning — a key lesson for any intervention operating within an emergency response.

## Data Availability

All data produced in the present study are available upon reasonable request to the authors.

## ACKNOWLEDGMENTS

This work would not have been possible without the support of PHI and KP staff that shared program information and documents with our team, including Marta Induni, Diane Royal, and Dana Williamson, and the CCTSI staff and stakeholders that participated in interviews.

