## Supplemental File for "Lessons in Implementing Complex Interventions in a Public Health Emergency: A Process Evaluation of the California Contact Tracing Support Initiative"

#### Supplemental Materials

##### Supplemental Tables

Supplemental Table 1. Inventory of files received from PHI and KP reviewed by Evaluation Team

| **Document Type** | **Total** |
| --- | --- |
| Training and Ongoing Development | 136 |
| KP-PHI leadership meeting notes and agenda | 78 |
| PHI and Evaluation Team Scoping discussion Agendas and Documents | 52 |
| CCTSI Implementation | 45 |
| KP Sponsors Weekly Update | 44 |
| Staff Recruitment | 40 |
| Workforce Development | 32 |
| Communications | 23 |
| Data Dictionaries | 6 |
| Grant agreement, Contracts, and Scopes of work | 6 |
| Data Procedures | 6 |
| Progress reports | 4 |
| Directories and Organizational Charts | 2 |
| **Total** | 474 |

Supplemental Table 2. Qualitative study participant characteristics

|  | n (%) | | |
| --- | --- | --- | --- |
| **Stakeholders:** | 24 (100) | | |
| Kaiser Permanente (KP) | 9 (38) | | |
| Public Health Institute (PHI) | 13 (54) | | |
| Other | 2 (8) | | |
|  | Total | Fresno | San Bernardino |
| **Micro-team (MT) members, n (row %):** | 23 (100) | 5 (22) | 18 (78) |
| Case investigator | 12 (52) | 3 (60) | 9 (50) |
| Resource coordinator | 4 (17) | 1 (20) | 3 (17) |
| Supervisor | 5 (22) | 1 (20) | 4 (22) |
| Team manager | 2 (9) | 0 (0) | 2 (11) |

Note: Column percentages are presented. Six MT members transferred their roles during the program period, but we reported the roles they maintained at the time of interview (for MT members still affiliated with the program) or the most recent roles (for MT members who left the program at the time of interview).

### Annexes

#### Program Design and Causal Assumptions of the California Contact Tracing Support Initiative (CCTSI)

While there are many reports of the outcomes and even processes of intervention implementation, too often there are not enough high quality and detailed accounts of the development and rationale of complex public health interventions for practitioners and researchers to effectively learn which elements were critical to program success or failure. Public health intervention research then needs to provide more detailed information on the rationale for the intervention model and the causal assumptions inherent in the program theory of change to better inform public health program planning and implementation. A detailed reporting of this contact tracing program is also useful for health services researchers seeking to understand key elements of integrated care pathways^19,20^.

##### Activities leading up to the establishment of CCTSI

The CCTSI program design elements, as well as the implicit causal assumptions underlying the program design, were derived in large part from Kaiser Permanente (KP) and Public Health Institute’s (PHI) pre-partnership experiences in San Jose, California and the Pacific Northwest.

###### The San Jose Pilot

In April-May of 2020, before Santa Clara County had started doing much contact tracing, clinicians at the KP medical center in San Jose, California decided to “bootstrap” some contact tracing work by calling KP patients who had tested positive for COVID-19 and performing their own case investigation. Although the clinicians who made calls were not patients’ primary care providers, members seemed reassured that they were part of the KP care team. Providers also connected KP members with social services to address issues such as food insecurity and transportation. This small-scale effort, which came to be called known colloquially as the “*pilot program*,” was anecdotally regarded as very successful, and became a catalyst for developing a clinically-integrated contact tracing model in partnership with KP, a trusted healthcare institution. KP believed this novel model would offer a more robust contact tracing program that could have a real impact on community spread of COVID-19. KP also envisioned replicating this contact tracing intervention in non-KP clinical settings, such as “*safety net*” institutions, or federally qualified health centers (FQHCs).

*"Initially, we [KP] designed this model with the intent that it was going to be more impactful in terms of timeliness, and breadth of services that are offered as well as being able to close the loop from the public health perspective on the contacts. And offer some support services as well." --KP Stakeholder*

###### PHI and the Pacific Northwest

Also in the Spring of 2020, PHI started working directly with county health departments in the Pacific Northwest (i.e., select county health departments in Oregon and Washington state) to provide support in the form of hiring, training, and managing staff to make contact tracing calls according to individual health department protocols. Although PHI implemented a different model of contact tracing in the Pacific Northwest (i.e., workflows and target priority populations were defined by local health jurisdictions metrics and priorities), the practical field experience gave them increased credibility with which to engage with KP on contact tracing support work in the US.

###### CCTSI Partnership

Recognizing that their recruiting, hiring, and onboarding processes for employees were slower than external agencies, KP sought an implementation partner to scale up the contact tracing intervention. KP’s decision to partner with PHI to implement the CCTSI was based on several considerations, including PHI’s organizational reputation, a history of inter-organizational collaboration, and a desire to find the partner best able to connect with their communities and make a community benefit impact. Several stakeholders from across KP and PHI noted that the success, both operationally and in terms of perceived impact, of PHI’s work in the Pacific Northwest during the Spring of 2020 was a key factor in KP’s decision to partner with PHI.

PHI was also able to employ the materials and best practices learned in the Pacific Northwest to CCTSI program design. For example, PHI demonstrated that they could hire people who were new to public health yet qualified for contact tracing by virtue of other skills, including mastery of non-English languages and lived experience – a practice which also aligned with KP’s commitment to improving contact tracing by increasing trust in callers.

*"We also knew that PHI was successfully partnering and bolstering the capabilities of local health jurisdictions in the Northwest in Washington State....we saw them as a partner who could actually do the hiring, who could do the training, and who could actually help to run the program if we helped to build it with them." -- KP Stakeholder*

*"We felt that they've [PHI have] already built some kind of models and some expertise in being able to hire people, train them, onboard them, so we thought they could be a good partner to get that work [contact tracing support work] going with them." – KP Stakeholder*

##### CCTSI program elements and causal assumptions

###### The California Contact Tracing Support Initiative

The CCTSI was primarily comprised of hiring, training and workforce development (WFD) activities, as well as case investigation and contact tracing and wraparound support services. Many of the contact tracing innovations came primarily from KP’s experience with the pilot, while many of the hiring, training, micro-team, and WFD elements of the program were borne out of PHI’s pre-partnership experiences. The program elements described below pertain primarily to the contact tracing intervention that was implemented in support of KP patients and their contacts:

###### Case Investigation and Contact Tracing

###### Trusted messenger

KP believed that their ability to leverage the existing trust of the patient-provider relationship with members was a key element of the perceived success of the San Jose pilot – an element that they were very committed to reproducing in the CCTSI. CCTSI call scripts were designed to describe the caller’s affiliation with KP to allow contact tracers to encourage case participation in contact tracing interviews and improve the quality of data collected during calls.

###### Direct clinical case dataflows

The contact tracing intervention was intended to function more quickly than the existing public health infrastructure by speeding up the process of transferring case test results/information to contact tracers (i.e., timeliness). KP surmised that embedding a contact tracing program into their integrated health system services would allow contact tracers to see SARS-CoV-2-positive test results as soon as they were entered into the KP electronic health records (EHR). Faster access to test data would allow contact tracers to start contact tracing with cases the same/next day after diagnosis as the KP providers did in the San Jose Pilot. Improved timeliness would help to slow disease transmission.

###### Wraparound services

Linking KP members with social services in the San Jose pilot led KP to hypothesize that people might be more willing to participate in a contact tracing program that provided other aid. To improve compliance with isolation or quarantine, CCTSI employed resource coordinators to inform beneficiaries about resources or services (e.g., food, household cleaning supplies, paying rent) for which they were eligible.

###### Closing the loop on clinical care

KP also wanted to link cases and contacts who were members back to KP clinical services. CCTSI work/call flow was designed to refer KP-members at the end of contact tracing calls to either an online e-Visit service or a Medical Assistant for test scheduling and other clinical care services. CCTSI also offered member-contacts the option of scheduling COVID-19 tests and daily monitoring calls to report symptoms during their quarantine period.

###### Micro-team structure

CCTSI contact tracing teams were called micro-teams and composed of 1 supervisor, 1 resource coordinator, 8-10 contact tracers, and later in the program, a Program Manager who managed coordination between many micro-teams. Micro-teams were intended to be effective for building team dynamics and adapting to the fast-paced context of pandemic response. Micro-team members were assigned to one local health jurisdiction each, however, could be moved around according to need.

###### Federally Qualified Health Centers (FQHCs)

To make services available to a larger subset of the population that would include non KP members, CCTSI intended to implement the program in select FQHCs in CA.

###### Hiring, Training, and Workforce Development

###### Impact hiring and cultural competency

CCTSI recruitment efforts targeted minority, low-income, and otherwise highly impacted neighborhoods to improve equity by increasing the employment rate. By removing the requirement for a bachelor’s degree and multiple interviews that were standard parts of PHI hiring protocols, CCTSI aimed to broaden their pool of applicants and give residents a unique opportunity for work that they otherwise might not have. CCTSI impact hiring policies and practices also placed high value on lived experiences, language skills, and cultural sensitivity as ways to facilitate trust during contact tracing calls.

*“The folks who envisioned [CCTSI] really thought ahead on that and saw a window of opportunity to recruit and inspire folks who might not have been on a public health pathway, but were desperately needed within public health, and built that in. And Kaiser supported it from the beginning."— CCTSI Stakeholder*

###### Comprehensive training curriculum

The CCTSI training curriculum was able to build on the work PHI did in the Pacific Northwest. It emphasized training in soft skills in addition to technical knowledge on contact tracing to help staff members build trust with call recipients, thus increasing the number of elicited contacts and the overall quality of investigation. Technical skills included contact tracing concepts, workflows and the use of data systems, while soft skill courses included active listening, showing empathy, and conducting motivational interviews:

*“We shadowed more experienced contact tracers, and they were able to show us the more nitty-gritty part of collecting information, how to go about writing your own, preparing your own script based on the original script, how to adhere to HIPAA laws, and some tips on how to save information and locate resources for that county. The shadowing portion of training was helpful in putting us into partners or teams to role-play and practice with each other, and [it taught us] how to prepare ourselves for questions that we might not see coming, or mentally prepare for reporting a positive result to a case.”– Micro-team Member*

###### Workforce Development Program

The CCTSI Workforce Development Program (WFD) was a set of activities intended to enhance the broader knowledge and interest of staff members in public health and related fields but were not directly related to completion of work-related training tasks. The goal was to build the capacity and skillset of the staff and introduce them to future public health job opportunities. The WFD Program focused on three different groups of staff: those new to the public health workforce, experienced employees with limited public health knowledge, and experienced public health practitioners. Courses included training and certifications through programs such as Coursera, mentoring opportunities, partnerships with UC Santa Cruz and Oakes College, tracking course completion as well as providing offboarding opportunities. Staff members were given designated paid time during the work week (8 hours) to engage in the WFD courses with the goal of increasing skillsets and interest in public health and “*bolstering and diversifying the public health workforce, especially in California*.” One micro-team member said of the WFD program:

*“Oh my God. It's the whole reason that I ended up back in school for the first time since 1988…So many of us took advantage of it. So, I took several courses, the Community Health Worker Program from El Sol was fantastic.”*

*“We wanted to act as a public health pipeline as well, to diversify those who are working in public health in this region and giving them hands-on experience to be able to really excel in those roles.” – PHI Stakeholder*

With equity and community benefit in mind, WFD activities were considered an “*equalizing strategy*” intended to influence staff beyond the life of the project. Other equalizing strategies included the remote work environment which eliminated the barrier of transportation and providing staff with equipment (e.g., laptops, headphones) and paid internet access for work.
